# Genetically Proxied Telomere Length but Not Epigenetic Aging Acceleration Causally Influences Healthspan: A Mendelian Randomization Study

**DOI:** 10.1101/2025.07.14.25331527

**Authors:** Bowen Feng, Robert Yang, Gabriella R Wang, Qiu Hu, Chuntao Zhao

## Abstract

**Objective:** To assess the causal effects of leukocyte telomere length (TL) and epigenetic age acceleration (EAA) on healthspan.

**Methods:** We performed two-sample Mendelian randomization (MR) analyses in accordance with STROBE-MR guidelines. Genetic instrumental variables (IVs) for TL and four EAA biomarkers (Hannum, GrimAge, PhenoAge, and intrinsic EAA) were derived from published genome-wide association study (GWAS) summary statistics involving up to 472,174 individuals for TL and approximately 35,000 individuals for each EAA biomarker. GWAS summary statistics for healthspan, defined as age at first diagnosis of any of eight major chronic conditions or death, were obtained from the UK Biobank (N=300,477 unrelated European-ancestry participants). The primary MR estimates were obtained using the inverse-variance weighted (IVW) method, complemented by various sensitivity analyses to assess pleiotropy, instrument heterogeneity, and robustness of causal inference. The strength of the IVs was evaluated using F-statistics, and causal directionality was validated using Steiger filtering.

**Results:** Genetically predicted longer TL was causally associated with extended healthspan (IVW β=0.106; 95% CI: 0.053–0.159; p=6.9×10^-5^). The association was robust across multiple sensitivity analyses, with no indication of directional pleiotropy (MR-Egger intercept p=0.47), no influential outliers identified by MR-PRESSO, and consistent causal direction confirmed by Steiger tests. In contrast, none of the four EAA biomarkers demonstrated convincing causal effects on health span (all IVW p-values >0.05), and results were inconsistent across sensitivity analyses, suggesting their role as correlates rather than causal determinants of healthy longevity.

**Conclusions:** This MR study provides robust evidence supporting a causal role of genetically determined telomere length in extending healthspan, while no such effect was observed for four commonly studied EAA biomarkers. These findings underscore the central role of telomere biology in healthy aging and indicate that telomere maintenance may represent a promising target for interventions aimed at delaying the onset of age-related diseases.

## Introduction

Aging is a complex biological process marked by progressive loss of physiological integrity, leading to impaired function and increased vulnerability to death. While lifespan, the total number of years an individual lives, has been the traditional focus of aging research, healthspan, defined as the duration of life spent in good health and free from major chronic diseases or disabilities, has emerged as a more meaningful target for improving the quality of aging populations (1, 2). Extending healthspan, rather than merely lifespan, addresses not just longevity but the burden of age-related morbidity and healthcare costs.

Unlike lifespan, which can be objectively measured, healthspan is a multidimensional construct that is often operationalized using composite indices of disease incidence, frailty, physical and cognitive function, and self-reported well-being. Multiple modifiable factors, such as smoking, low socioeconomic status, obesity, and chronic inflammation, have been found to be associated with a reduced healthspan. For instance, smoking is linked to increased systemic inflammation and elevated levels of C-reactive protein (CRP), contributing to various chronic diseases. A previous study using data from the Heath and Retirement Study (HRS) found that male smokers have over two years less healthy life remaining compared to never smokers, and female smokers have 1.66 fewer such years (3). Low socioeconomic status is associated with heightened inflammation (4) and a higher risk of chronic conditions (5), which can shorten healthspan (6). Obesity contributes to chronic low-grade inflammation, known as meta-inflammation (7), which plays a role in the development of age-related diseases, such as type 2 diabetes (8), nonalcoholic fatty liver disease (NAFLD) (9), and atherosclerosis (10). Collectively, these factors contribute to the earlier onset of chronic diseases, thereby reducing the duration of life spent in good health.

Genome-wide association studies (GWAS) and epigenetic analyses have begun to uncover the genetic and epigenetic architecture of healthspan, identifying loci linked to immune regulation, metabolism, and inflammation. For instance, a previous study identified 12 genetic loci associated with healthspan using a large-scale GWAS in the UK Biobank cohort (11), while a recent study leveraging UK Biobank data of over 241,000 individual reported 13 novel DNA methylation markers that predict healthspan termination (12). Another study combining GWAS of three ageing phenotypes, including lived in good health (healthspan), total years lived (lifespan), and survival until an exceptional old age (longevity), identified 10 genomic loci that influence all three ageing phenotypes (13). A more recent study constructed principal components (PCs) using six human aging traits, including healthspan. Using two-sample Mendelian randomization (MR), it provided robust evidence for a detrimental effect of blood levels of apolipoprotein(a) and vascular cell adhesion molecule 1 on aging-GIP1, the first PC capturing both length of life and indices of mental and physical wellbeing (14). These findings highlight shared pathways with multiple age-related traits and diseases, including metabolic syndrome, cardiovascular disease, and dementia.

Among the molecular markers of aging, telomere length and epigenetic age acceleration (EAA) have been widely studied. Telomeres, repetitive DNA sequences that cap the ends of chromosomes, shorten with each cell division and are considered hallmarks of cellular aging (15). Shorter leukocyte telomere length (TL) has been associated with numerous age-related diseases, including cardiovascular disease, type 2 diabetes, and all-cause mortality (16–18). Separately, epigenetic clocks, which estimate biological age based on DNA methylation patterns, have emerged as powerful predictors of age-related outcomes (19). Clocks such as Hannum (20), GrimAge (21), PhenoAge (22), and intrinsic epigenetic age acceleration (IEAA) (19, 23) have shown correlations with frailty (24), chronic diseases (25), mortality (26), lifespan, or healthspan (22) in observational studies.

However, a key limitation of prior work is the challenge of disentangling causality from correlation. Observational associations are susceptible to confounding, reverse causation, and measurement bias. MR, an analytical approach using genetic variants as instrumental variables, can help overcome these limitations by inferring causality under specific assumptions (27). Although MR has been applied to assess the causal role of telomere length in individual diseases (28, 29), and recently, to a limited extent, with EAA measures (30), the comparative causal relevance of these aging biomarkers on healthspan, a holistic measure of aging, remains underexplored.

In this study, we used two-sample MR to investigate whether genetically proxied telomere length and four established epigenetic aging biomarkers (Hannum, GrimAge, PhenoAge, and IEAA) exert causal effects on healthspan. We leveraged publicly available GWAS summary statistics for each exposure and outcome and applied a suite of robust MR sensitivity analyses to account for pleiotropy, heterogeneity, and potential violations of instrument validity. Our findings provide novel evidence that telomere length—but not epigenetic age acceleration—is causally associated with increased healthspan, suggesting that telomere maintenance may be a key mechanism for extending healthy longevity.

## Methods

We performed a two-sample MR study to evaluate the causal relationships between leukocyte telomere length and four EAA biomarkers, including Hannum, GrimAge, PhenoAge, and IEAA, and healthspan. Analyses were conducted in accordance with the STROBE-MR reporting guidelines (31).

### Data Sources and Study Design

This study utilized publicly available genome-wide association study (GWAS) summary statistics from European ancestry cohorts. GWAS data for leukocyte telomere length were obtained from a meta-analysis conducted by Codd et al. (32), involving up to 472,174 participants from the UK Biobank and other population-based studies. Telomere length was measured in leukocytes using a validated quantitative polymerase chain reaction (qPCR)-based method to estimate the telomere repeat copy number relative to a single-copy gene (T/S ratio) (32).

Healthspan data were derived from Zenin et al. (11), based on 300,477 participants from the UK Biobank. Healthspan was operationalized as the age at first diagnosis of any of eight chronic diseases: cancer, congestive heart failure, myocardial infarction, chronic obstructive pulmonary disease, stroke, dementia, diabetes, or death. Disease onset was determined through linked hospital records and national death registries rather than self-report, ensuring consistency and clinical validity.

Summary statistics for the four EAA biomarkers were obtained from McCartney et al. (33), which conducted GWAS meta-analyses of Hannum, GrimAge, PhenoAge, and IEAA acceleration in up to 34,710 European ancestry individuals across 24 cohorts. All four EAA measures were constructed using DNA methylation data derived from whole blood samples, primarily assayed using the Illumina 450K and EPIC arrays. Each biomarker was calculated using previously established epigenetic clock algorithms: Hannum and Horvath’s clocks were based on penalized regression models using methylation levels at selected CpG sites (20), while GrimAge and PhenoAge integrated methylation surrogates for plasma proteins and clinical parameters associated with aging and morbidity (21, 22). Residuals from regressing predicted epigenetic age on chronological age were used to calculate IEAA (19, 23).

All GWAS studies included appropriate quality control procedures and adjustments for age, sex, and principal components. Ethical approval and informed consent were obtained in the original studies.

### Quality Control of the Summary Data

Prior to MR analysis, we implemented several quality control (QC) steps for each GWAS dataset to ensure validity and consistency. Specifically for the telomere length dataset, we retained only variants with standard rsID identifiers, removed SNPs with missing values in key variables (e.g., p-values, beta, standard error, allele frequency), filtered to include only biallelic SNPs (i.e., single-letter alleles), and excluded ambiguous strand SNPs (A/T or C/G). Variants with out-of-bounds allele frequencies (≤0 or ≥1) were discarded.

Additionally, duplicated SNP identifiers were removed. Equivalent QC procedures were applied to the other exposure and the outcome datasets to harmonize data inputs for downstream MR analysis.

### Selection and Harmonization of Genetic Instruments

For each exposure, independent SNPs were selected at genome-wide significance (p < 5 × 10□□) and clumped using a linkage disequilibrium threshold of r² < 0.001 within a 10 Mb window, based on the 1000 Genomes Project European reference panel. For telomere length, where the number of genome-wide significant variants was large, we applied a more stringent Bonferroni threshold based on the total number of SNPs tested to minimize the inclusion of weak or pleiotropic instruments.

Data harmonization was performed using the harmonise_data function in the TwoSampleMR R package to ensure consistent alignment of effect alleles. Palindromic SNPs with intermediate allele frequency were excluded. The analytical workflow for the MR analysis investigating the causal effect of telomere length on healthspan is illustrated in **Figure 1**. A similar workflow was applied to the other MR analyses.

**Figure 1.**
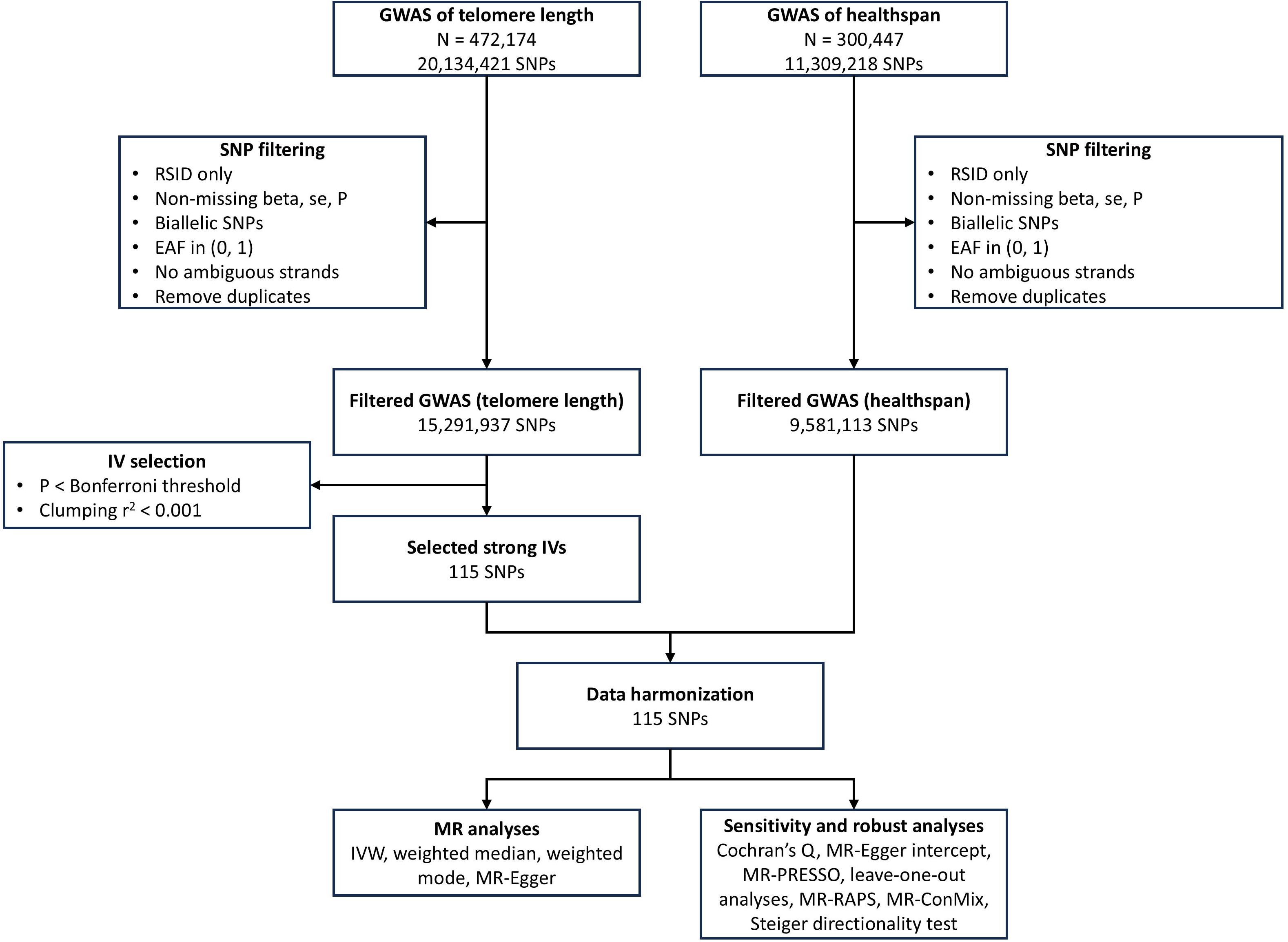
Analytical workflow for MR analysis assessing the causal effect of telomere length on healthspan. The workflow includes selection of IVs from GWAS, harmonization of exposure and outcome datasets, MR analysis using multiple methods (e.g., inverse-variance weighted and weighted median), and sensitivity analyses to evaluate robustness. This workflow was similarly applied to other MR analyses in the study. MR, Mendelian randomization; GWAS, genome-wide association study; IV, instrumental variable

### Mendelian Randomization Assumptions

MR analyses rely on three key assumptions: (1) the genetic variants are strongly associated with the exposure (relevance), (2) the genetic variants are independent of confounders of the exposure-outcome relationship (independence), and (3) the genetic variants influence the outcome solely through the exposure (exclusion restriction). These assumptions were assessed through multiple sensitivity analyses described below.

### Statistical Analyses

Causal estimates were primarily derived using the inverse-variance weighted (IVW) method under a multiplicative random-effects model, along with other estimators, including weighted median, weighted mode, and MR Egger. We calculated F-statistics (F = β² / SE²) to assess instrument strength, with F > 10 considered indicative of sufficient strength. All exposure and outcome variables were analyzed on a continuous scale using standardized effect estimates.

No individual-level data were applicable as the study used summary-level data. SNPs that could not be harmonized across exposure and outcome datasets were excluded. Multiple testing adjustments were not applied formally, but we interpreted results conservatively, given the inclusion of five exposures.

### Sensitivity Analyses

To evaluate the robustness of our MR findings and assess the validity of core instrumental variable assumptions, we conducted a comprehensive suite of sensitivity analyses. Heterogeneity across SNP-specific causal estimates was examined using Cochran’s Q statistic. To detect potential directional pleiotropy, we employed the MR-Egger intercept test. We also applied the MR-PRESSO framework to identify and correct for horizontal pleiotropy through global and distortion tests (34). To evaluate the influence of individual SNPs on the overall estimate, we performed leave-one-out analyses. Furthermore, we explored the consistency of our findings by comparing results across multiple robust MR estimators, including MR-RAPS (35) and the contamination mixture model (MR-ConMix) (36). To assess whether the direction of causality was consistent with our hypothesis, we performed the Steiger directionality test. This test evaluates whether the genetic variants explain more variance in the exposure (e.g., telomere length) than in the outcome (e.g., healthspan), helping to confirm that the exposure-outcome relationship is not driven by reverse causation (37).

### Software and Reproducibility

All analyses were performed using R version 4.4.3 (https://www.R-project.org/). Core packages included TwoSampleMR, MRPRESSO, mr.raps, and MendelianRandomization. This study was not preregistered.

## Results

### Descriptive Data

The MR analyses were conducted using summary-level GWAS data from large-scale studies on participants of European ancestry. The exposure data consisted of leukocyte telomere length (up to 472,174 individuals) (32) and four EAA biomarkers, including Hannum, GrimAge, PhenoAge, and IEAA, with up to 34,710 individuals (33). The outcome data for health span included 300,477 individuals from the UK Biobank (11) (**Figure 1**). Detailed information of the data is summarized in **Table 1**.

**Table 1.** Basic information of the GWAS summary data.

|  | **TL** | **Hannum** | **GrimAge** | **PhenoAge** | **IEAA** | **Heathspan** |
| --- | --- | --- | --- | --- | --- | --- |
| # of subjects | 472,174 | 34,710 | 34,710 | 34,710 | 34,710 | 300,447 |
| # of genetic variants | 20,134,421 | 6,419,390 | 6,421,697 | 6,421,578 | 6,421,541 | 11,309,218 |
| Age | 56.5 (8.1) | 53.2 (15.4) | 53.2 (15.4) | 53.2 (15.4) | 53.2 (15.4) | 37-73 |
| Sex male, n (%) | 216,187 (45.8%) | 14,242 (41.0%) | 14,242 (41.0%) | 14,242 (41.0%) | 14,242 (41.0%) | 137,469 (45.8%) |

Age was presented as mean (SD) or range.
GWAS, genome-wide association study; SD, standard deviation; IEAA, intrinsic epigenetic age acceleration; TL, telomere length

### Telomere Length and Healthspan

We identified 115 independent SNPs as IVs for leukocyte telomere length (**Table S1**). The IVW analysis indicated a significant positive causal effect of genetically proxied telomere length on healthspan (beta = 0.106, SE = 0.027, p = 6.86 × 10^-5^). Consistent results were observed across multiple robust MR methods, including the weighted median (beta = 0.151, SE = 0.034, p = 9.93 × 10^-6^), weighted mode (beta = 0.163, SE = 0.042, p = 1.57 × 10^-4^), and MR-Egger regression (beta = 0.215, SE = 0.047, p = 1.13 × 10^-5^) (**Figure 2**).

**Figure 2.**
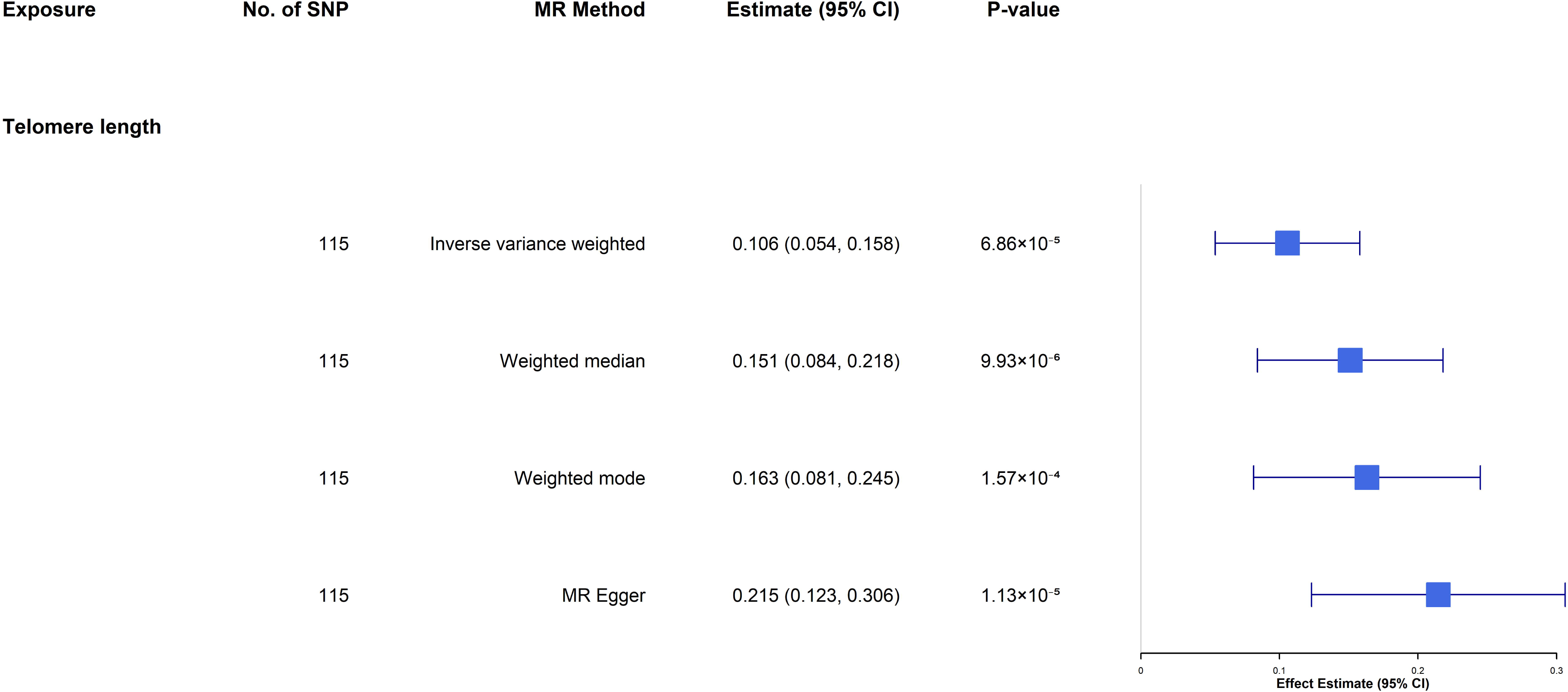
Forest plot of MR estimates of the causal effect of genetically proxied telomere length on healthspan. The forest plots summarize the causal effect estimates of telomere length on healthspan. We used four MR methods: IVW, weighted median, weighted mode, and MR Egger regression. Each method is represented by a point estimate and a 95% CI, along with the corresponding p-values. MR, Mendelian randomization; IVW, inverse-variance weighted; IEAA, intrinsic epigenetic age acceleration; CI, confidence interval; SNP, single-nucleotide polymorphism

The MR-PRESSO global test detected significant horizontal pleiotropy (RSS_obs_ = 201.67, p < 0.0003). After removing outlier SNPs, the causal effect remained significant (MR-PRESSO corrected estimate: beta = 0.102, SE = 0.026, p = 1.27 × 10^-4^). We observed a robust and consistent positive causal effect of telomere length on healthspan across multiple MR approaches. MR-RAPS produced consistent estimates (β = 0.11, SE = 0.021), and the MR-ConMix estimated a causal effect of β = 0.16 (95% CI: 0.10–0.23, p = 1.22 × 10^-4^), further supporting the validity of the association. Radial MR identified 14 outlier SNPs, the removal of which slightly increased the IVW effect estimate (β = 0.13, SE = 0.022, p = 8.6 × 10^-9^) and resolved heterogeneity (Q = 100.3, p = 0.47). No evidence of directional pleiotropy was detected after outlier removal (MR-Egger intercept p = 0.24), and leave-one-out analysis confirmed that the results were not driven by single influential SNPs (**Figure S1**). The Steiger directionality test supported the hypothesized causal direction from telomere length to health span (p < 1 × 10^-4^). Together, these findings provide strong and consistent evidence for a causal role of telomere length in promoting healthspan.

#### Epigenetic Age Acceleration Biomarkers and Healthspan

We assessed the potential causal effects of four commonly used EAA biomarkers on healthspan using two-sample MR. Across all four measures, we found no strong or consistent evidence supporting a causal relationship.

For the Hannum clock (8 SNPs as IV, **Table S2**), all MR estimators produced null findings (**Figure S2**). The IVW estimate was β = 0.007 (SE = 0.012, p = 0.540), and MR-Egger similarly yielded a non-significant association (β = 0.033, SE = 0.055, p = 0.567). While the MR-PRESSO global test indicated marginal evidence of heterogeneity (p = 0.052), no outliers were corrected, and the raw causal estimate remained non-significant.

In the GrimAge analysis (4 SNPs as IV, **Table S3**), the IVW method yielded a nominally significant estimate (β = 0.032, SE = 0.014, p = 0.018); however, this association was not corroborated by the weighted median (p = 0.083) or MR-Egger (p = 0.491), suggesting a lack of robustness (**Figure S2**). MR-PRESSO found no evidence of pleiotropy, and outlier correction did not materially alter the estimate.

For PhenoAge (11 SNPs as IV, **Table S4**), none of the MR methods showed significant associations (**Figure S2**), with IVW yielding β = 0.006 (SE = 0.005, p = 0.237). Likewise, the MR-Egger and weighted median results were non-significant, and MR-PRESSO did not identify any influential outliers or pleiotropy.

Finally, the analysis of IEAA (24 SNPs as IV, **Table S5**) also returned consistently null results across all MR approaches (**Figure S2**). The IVW estimate was β = 0.004 (SE = 0.004, p = 0.381), with no evidence of heterogeneity (MR-PRESSO p = 0.946) or pleiotropy, and the results remained stable across alternative MR methods.

#### Assessment of Assumptions

To evaluate the validity of the core MR assumptions, we undertook several diagnostic and sensitivity analyses. The relevance assumption, which requires that the selected genetic instruments are strongly associated with the exposure, was supported by consistently high F-statistics across SNPs (**Tables S1-S5**). For telomere length (**Table S1**), the mean F-statistic was 125.3 (range: 35.1–1350.3), indicating that all instruments were strongly predictive of the exposure (F > 10), thus minimizing the risk of weak instrument bias.

The independence assumption posits that the genetic variants are not associated with confounders of the exposure-outcome relationship. While this assumption cannot be directly tested using summary-level data, we mitigated potential violations by restricting to genome-wide significant variants (Bonferroni threshold for telomere length or p < 5 × 10^-8^ for EAAs), performing LD clumping (r² < 0.001, 10 Mb window), and restricting analyses to individuals of European ancestry to minimize population stratification. All source GWAS were adjusted for key covariates, including age, sex, and population structure via principal components.

To assess the assumption of exclusion restriction, we conducted multiple pleiotropy-detection analyses. For the telomere length–healthspan analysis, MR-Egger regression revealed no evidence of directional pleiotropy after outlier removal (intercept = –0.0013, p = 0.24). The MR-PRESSO global test detected significant horizontal pleiotropy (RSS_obs_ = 201.67, p < 0.0003). After removing outlier SNPs, the causal effect remained significant (MR-PRESSO corrected estimate: beta = 0.102, SE = 0.026, p = 1.27 × 10^-4^). The heterogeneity was substantially reduced. Specifically, Cochran’s Q statistics for IVW and MR-Egger dropped to 100.3 (p = 0.47) and 98.9 (p = 0.48), respectively, indicating low heterogeneity among the remaining instruments. These results were further supported by Radial MR, which identified the same 14 outlier variants and produced highly consistent estimates (β = 0.125, SE = 0.022, p = 8.6 × 10^-9^).

For the EAA biomarkers, which yielded null results, heterogeneity and pleiotropy statistics did not indicate significant violations of MR assumptions. MR-PRESSO global tests for all four EAA clocks were non-significant (p > 0.05), and MR-Egger intercepts showed no evidence of directional pleiotropy. Therefore, while the absence of causal effects limits the interpretability of assumption diagnostics, no evidence suggested that violations of MR assumptions explain the null findings.

## Discussion

In this two-sample MR study, we systematically evaluated the causal relationships of genetically predicted leukocyte telomere length and four commonly used EAA biomarkers with healthspan. Consistent with our primary hypothesis, we found robust evidence that longer genetically proxied telomere length causally increases healthspan. In contrast, we found no consistent evidence for a causal effect of any of the four EAA biomarkers on healthspan. These findings suggest that among the key molecular biomarkers of biological aging, telomere length may play a uniquely causal role in promoting a longer period of life free from major chronic diseases. The null results for epigenetic clocks also emphasize the importance of distinguishing between biomarkers that reflect aging and those that drive it.

Telomeres, the repetitive nucleotide sequences (TTAGGG) at the ends of eukaryotic chromosomes, play a critical role in preserving genome stability by protecting chromosome ends from degradation, end-to-end fusions, and inappropriate DNA damage responses (38). During somatic cell division, telomeres progressively shorten due to the end-replication problem and oxidative stress, eventually triggering cellular senescence or apoptosis once a critical threshold is reached (39). This process, often referred to as the “mitotic clock,” is central to the aging of proliferative tissues such as the hematopoietic system, skin, and gastrointestinal epithelium (40). Telomere attrition is not merely a passive marker of aging but actively contributes to age-related pathologies by promoting stem cell exhaustion, mitochondrial dysfunction, increased inflammation (inflammaging), and impaired tissue regeneration (41, 42). The protective capping of telomeres by the shelterin complex, which includes proteins such as TRF1, TRF2, POT1, and TIN2, is essential for suppressing the DNA damage response and maintaining genomic integrity (43). Disruption of shelterin function or critically short telomeres can activate p53-mediated pathways that limit cell proliferation and tissue maintenance. Our results suggest that genetically proxied telomere length may play a causal role in promoting healthspan.

EAA biomarkers, such as GrimAge and PhenoAge, have also been found to be strongly associated with morbidity and all-cause mortality in prospective cohort studies (21, 22, 26), yet our null MR findings suggest that these associations may be non-causal and driven by confounding or reverse causality. One possible explanation lies in the origin and design of these biomarkers. All four EAA biomarkers included in our study were developed using DNA methylation data derived exclusively from whole blood. Given the known tissue specificity of epigenetic aging processes (44), they may not fully capture epigenetic aging processes in disease-relevant tissues such as the brain, heart, or kidney. Consequently, EAAs may be limited in their ability to detect causal pathways for systemic aging or organ-specific morbidity. Furthermore, although EAA traits are statistically robust predictors of age and morbidity, the underlying genetic architecture of these clocks explains only a modest proportion of their variance [8]. This limited heritability reduces instrument strength in MR analyses, potentially undermining the statistical power to identify small but meaningful causal effects. Additionally, EAAs may function as distinct biomarkers that reflect different underlying “hallmarks” or “pillars” of aging (45). The lack of causal association with healthspan in our study may thus reflect the fact that EAAs are downstream correlates of biological aging, shaped by complex interactions between genetic, epigenetic, and environmental factors rather than singular upstream determinants.

Our study has several limitations that warrant consideration. First, the accuracy of telomere length estimation is a known concern. The GWAS of leukocyte telomere length used in our analysis utilized both qPCR and WGS-based methods in UK Biobank participants (32). While these techniques are widely adopted and cost-effective, qPCR in particular is susceptible to inter-assay variability, batch effects, and signal-to-noise ratio issues, which may introduce non-differential measurement error (46). Such measurement imprecision can bias effect estimates toward the null, potentially attenuating observed associations in both GWAS and downstream MR analyses (47). Second, there is a potential sample overlap between the GWAS datasets for the exposures (e.g., telomere length) and the outcome (healthspan), as both include participants from the UK Biobank. Sample overlap can introduce weak instrument bias in two-sample MR settings, especially when the genetic instruments explain a modest proportion of exposure variance. In such cases, the MR estimate may be biased toward the confounded observational association, leading to overestimation of causal effects (48). Although our instruments for telomere length demonstrated strong F-statistics and we performed clumping to ensure SNP independence, this limitation cannot be fully excluded. Third, our analysis assumes that the instrumental variables satisfy core MR assumptions, particularly the exclusion restriction assumption, i.e., the genetic variants influence the outcome solely through the exposure. We employed MR-Egger regression and MR-PRESSO global and distortion tests to detect and correct for horizontal pleiotropy, which showed no significant evidence of bias. Nonetheless, these methods are known to have limited power, especially with a modest number of variants or when the InSIDE (Instrument Strength Independent of Direct Effect) assumption is violated (34, 49). Thus, undetected pleiotropic effects may still have influenced our estimates. In terms of generalizability, our findings are most relevant to individuals of European ancestry, as all GWAS datasets used were based primarily on European-descent populations. Applicability to other ancestral groups remains uncertain, and trans-ethnic validation is needed. Finally, our study captures lifelong, genetically determined differences in telomere length and EAA; it does not account for short-term or environmentally modifiable influences on these aging biomarkers. Therefore, extrapolating our findings to the effects of behavioral or pharmacologic interventions requires caution.

In conclusion, this MR study provides robust evidence supporting a causal role of genetically determined telomere length in extending healthspan, while no such effect was observed for four commonly studied epigenetic age acceleration biomarkers. These findings underscore the primacy of telomere biology in healthy aging and suggest that telomere maintenance may be a promising target for interventions aimed at delaying the onset of age-related diseases. Future studies should explore the causal relevance of telomere dynamics across diverse populations and tissues and examine whether modifiable lifestyle or pharmacologic interventions can replicate the genetic effects observed here.

## Declarations

## Ethical Approval

This study used only publicly available data. As a result, ethical approval and consent to participate are not needed.

## Authors’ Contributions

C.Z. designed the study. B.F., R.Y., G.R.W., and R.H. analyzed data and performed data interpretation. B.F. and C.Z. wrote the initial draft, and R.Y., G.R.W., and R.H. contributed to writing subsequent versions of the manuscript. The manuscript was written with contributions from all authors. All authors approved the final version before submission.

## Data Availability

All data generated or analyzed during this study are publicly available as specified in the methods section of this paper. Specifically, the GWAS summarized data for telomere length can be downloaded at https://figshare.com/s/caa99dc0f76d62990195?file=28414941, the GWAS summarized data for the four epigenetic clocks can be downloaded at https://datashare.ed.ac.uk/handle/10283/3645, and the GWAS summary data for healthspan can be downloaded at https://zenodo.org/records/1302861.

## Consent to participate

NA.

## Consent to publish

NA.

## Figure legends

**Figure S1. Leave-One-Out sensitivity analysis for the effect of genetically proxied telomere length on healthspan.**

This leave-one-out MR sensitivity analysis evaluates the robustness of the estimated causal effect of genetically proxied telomere length on healthspan. Each black dot and horizontal line represent the causal estimate and its 95% confidence interval obtained by sequentially removing one SNP from the analysis. The red dot and line at the bottom denote the overall IVW estimate using all SNPs.

The stability of the effect estimate across all iterations suggests that no single SNP unduly influences the overall association, indicating that the result is not driven by outlier instruments.

MR, Mendelian randomization; SNP: single-nucleotide polymorphism; IVW, inverse-variance weighted

**Figure S2. Forest plots of MR estimates of the causal effect of epigenetic age acceleration on healthspan.**

The forest plots summarize the causal effect estimates of four epigenetic age acceleration measures, including Hannum age, GrimAge, PhenoAge, and IEAA, on healthspan. We used four MR methods: IVW, weighted median, weighted mode, and MR Egger regression. Each method is represented by a point estimate and a 95% CI, along with the corresponding p-values.

MR, Mendelian randomization; IVW, inverse-variance weighted; IEAA, intrinsic epigenetic age acceleration; CI, confidence interval; SNP, single-nucleotide polymorphism

