## Supplementary figures and images for "Genetically Proxied Telomere Length but Not Epigenetic Aging Acceleration Causally Influences Healthspan: A Mendelian Randomization Study"

### Supplementary Figure 1

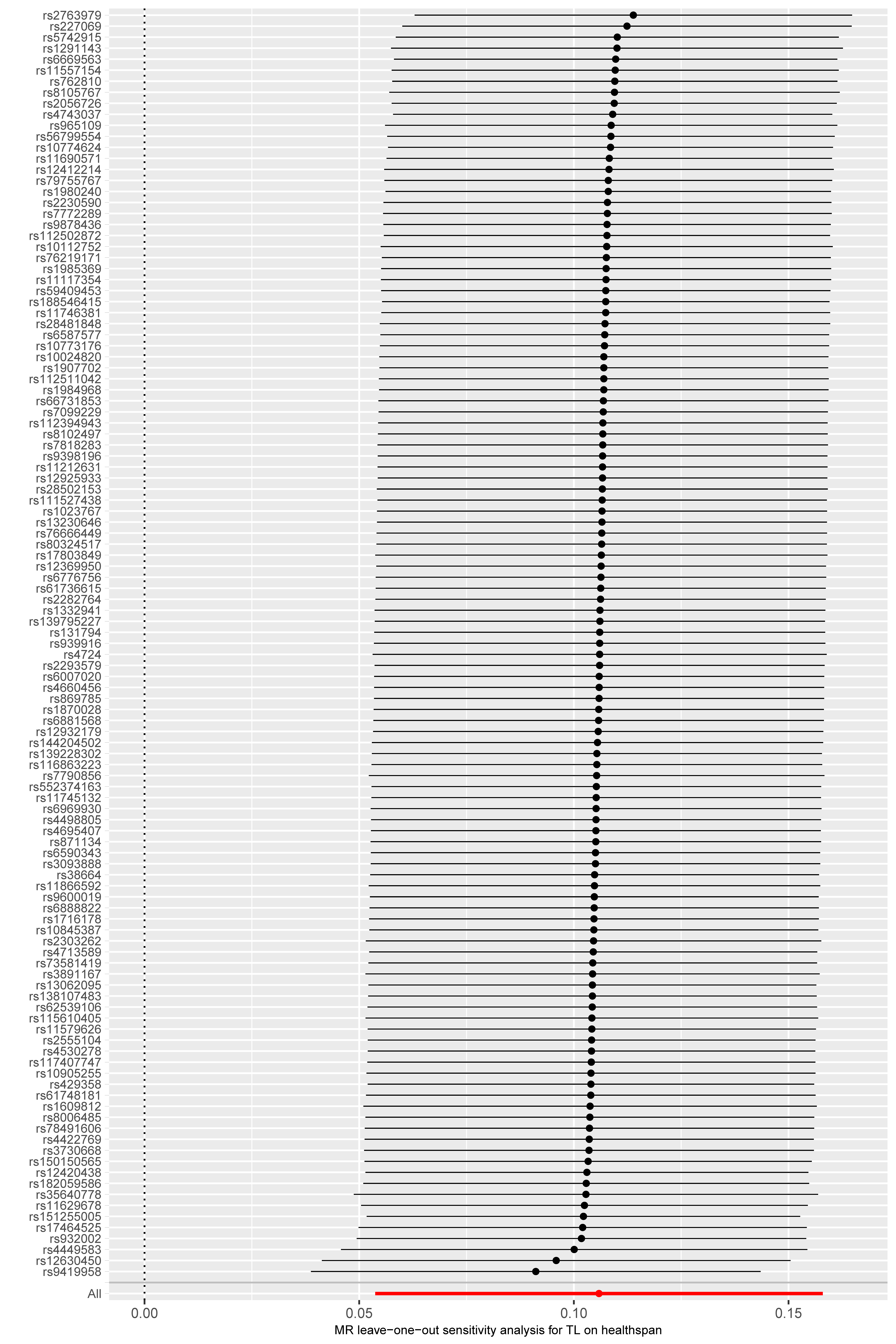

### Supplementary Figure 2

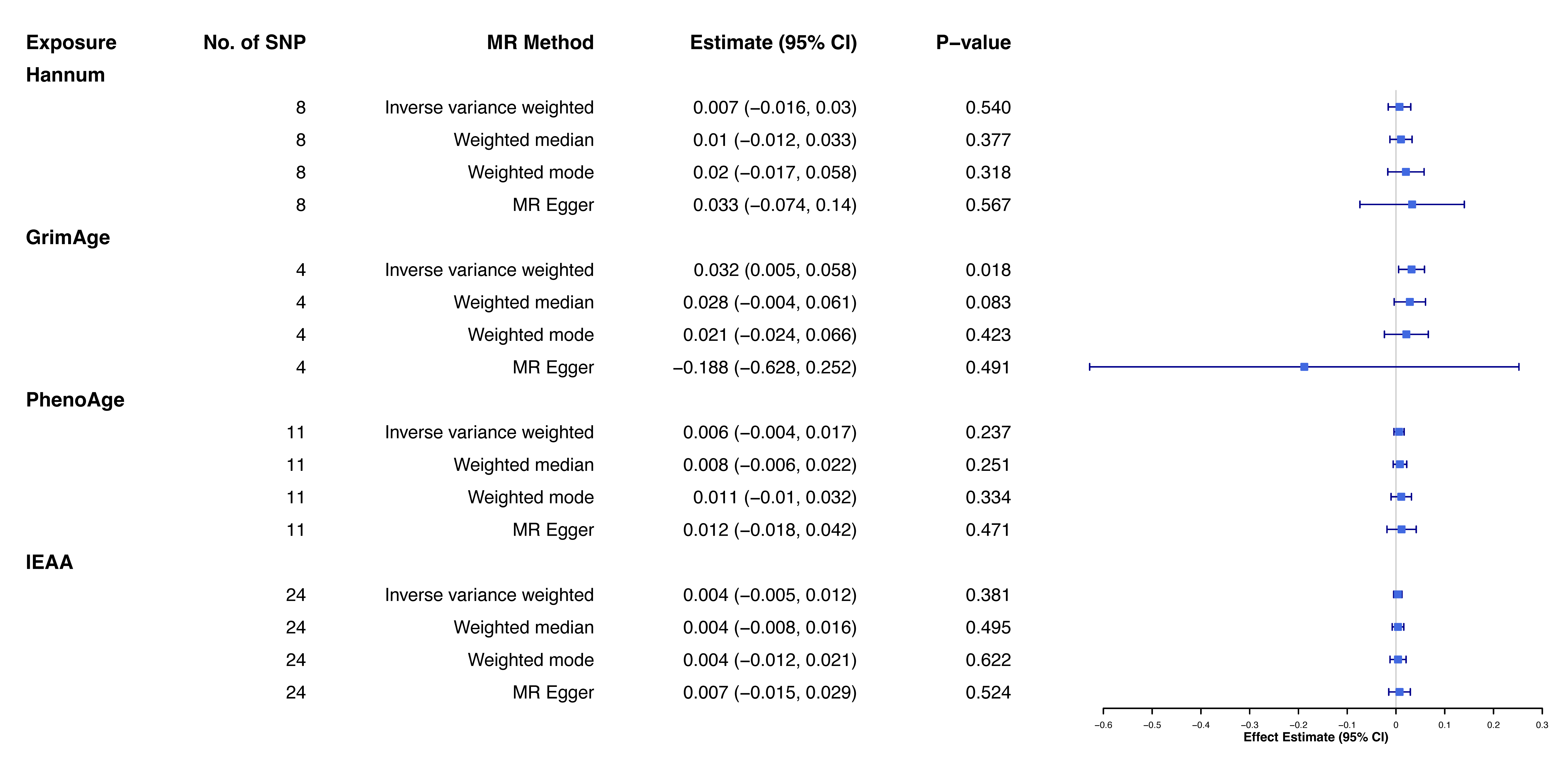
